# An FDA-Approved Tenofovir Alafenamide-Based Antiretroviral Therapy Reduces Biological Age in Healthy Adults: First Human Proof-of-Concept for Retrotransposon-Targeted Gerotherapeutics

**DOI:** 10.64898/2026.03.23.26349105

**Authors:** Peter L. Anderson, Alina PS Pang, Ryan P. Coyle, Johannes Schlachetzki, Anthony J.A. Molina, Lane Bushman, Julio Aguado, Blake Hill, Albert Y. Liu, Kristina M. Brooks, Kristine M. Erlandson, Michael J. Corley

## Abstract

Nucleos(t)ide reverse transcriptase inhibitors (NRTIs) used for HIV treatment and pre-exposure prophylaxis have been proposed as gerotherapeutics based on their capacity to suppress age-associated retrotransposon activity. However, evidence in humans is currently lacking. Here we evaluated DNA methylation–based measures of biological aging in healthy people without HIV (aged 18–50) using samples from two separate randomized, directly observed dosing pharmacokinetic studies of FDA-approved NRTI regimens containing emtricitabine–tenofovir-alafenamide (FTC/TAF;200 mg/25 mg) or FTC-tenofovir-disoproxil fumarate (FTC/TDF; 200 mg/300 mg) for 12 weeks. In the FTC/TAF study (N=36), epigenetic aging measures based on DNA methylation (DNAm) profiling decreased over follow-up, including DunedinPACE (−0.061, p=0.019) and PhenoAge (−6.33, p=0.008), with concordant reductions (p<0.05) across additional systems-specific epigenetic clocks including those estimating brain aging. DNAm–based proxies of inflammatory biomarkers also declined, with significant reductions in epigenetic IL-6 (−0.058, p=0.029) and a trend toward reduced C-reactive protein (−0.231, p=0.059). In contrast, the FTC/TDF study (N=43) showed no significant changes across epigenetic clocks and proxies. These findings are consistent with TAF’s more favorable cellular pharmacology compared with TDF and support gerotherapeutic effects of FTC/TAF. Prospective placebo-controlled studies are warranted that integrate clinical pharmacology, direct transposable element readouts, and prespecified geroscience and DNA methylation–based aging endpoints.

## INTRODUCTION

Nearly half of the human genome (∼45%) is composed of transposable elements (TEs)^1^. Aging is accompanied by a progressive erosion of epigenetic silencing that permits the transcriptional reactivation of these TEs, particularly retrotransposons such as LINE-1 and endogenous retroviruses^2–10^. In young somatic cells, these elements are maintained in a transcriptionally inert state by DNA methylation, heterochromatin, and KRAB-ZFP/KAP1 surveillance. However, with age the fidelity of these mechanisms declines, and retrotransposon-derived transcripts and cytoplasmic DNA accumulate^2^. This age-dependent retroelement reactivation is now recognized as a proximal driver of biological aging hallmarks including a senescence-associated secretory phenotype (SASP) and age-related tissue dysfunction^11–15^.

The dependence of retroelements on reverse transcription has made nucleos(t)ide reverse transcriptase inhibitors (NRTIs), which were developed and licensed for HIV treatment and prevention, attractive candidate gerotherapeutics^16^. For instance, a retrospective analysis of longitudinal aging intervention studies identified antiretroviral therapy as one of the most consistent interventions associated with reductions across 16 epigenetic clocks^17^. Early mechanistic work showed that multiple NRTIs including 3TC (lamivudine), tenofovir disoproxil fumarate (TDF), stavudine, and zidovudine can directly suppress human LINE-1 retrotransposition in cell-based reporter systems^18^. Consistent with this, 3TC (lamivudine) blunted LINE-1 cDNA–triggered type I interferon signaling and components of the SASP in senescent human cells and reduced age-associated inflammatory signatures across multiple tissues in aged mice^11^. In a complementary genetic model, NRTI treatment inhibited the expansion of LINE-1 DNA/cytoplasmic cDNA, rescued aberrant type I interferon responses, and improved health and survival in SIRT6-deficient mice that exhibit accelerated aging-like phenotypes^19^. Beyond LINE-1, NRTI treatment antagonized endogenous retrovirus (ERV)–linked senescence and tissue aging programs^12^. Prior work has also shown that FDA-approved NRTIs inhibited HERV-K reverse transcriptase activity and reduced HERV-K infection/production in a cell-based pseudotype assay^20^. Finally, recent work in neurodegeneration models and clinical trials reports that 3TC (lamivudine) may reduce age-associated brain inflammatory/transcriptomic signatures extending potential NRTI effects to brain aging^21,22^.

A critical question, however, is whether the retrotransposon-suppressive and anti-inflammatory effects of NRTIs translate into measurable changes in biological aging in healthy adults without HIV or age-related disease. Equally important is whether regimen-specific pharmacokinetics influence any such effect. The two globally prescribed emtricitabine–tenofovir formulations for HIV pre-exposure prophylaxis, emtricitabine/tenofovir alafenamide (FTC/TAF; Descovy) and emtricitabine/tenofovir disoproxil fumarate (FTC/TDF; Truvada), share the same close analog to 3TC (lamivudine) (emtricitabine) but differ fundamentally in their tenofovir prodrug. TAF circulates systemically, delivering lipophilic prodrug to target cells where tenofovir is liberated, thereby achieving substantially higher active tenofovir-diphosphate (TFV-DP) concentrations across immune cell tissues with lower systemic tenofovir exposures. Conversely, TDF is rapidly metabolized to tenofovir which is ionized and poorly penetrates cells leading to lower active TFV-DP in immune cells with higher plasma tenofovir ^23,24^. Since cells are the compartment where L1 de-repression drives cGAS–STING-mediated inflammation and the cellular source for DNA methylation profiling, this pharmacokinetic divergence between TAF and TDF provides a comparative setting to test whether intracellular drug exposure is a relevant factor of any gerotherapeutic signal.

Here we leveraged longitudinal specimens from two separate randomized, directly observed dosing pharmacokinetic studies, FTC/TAF (NCT02962739^25^) and FTC/TDF (NCT02022657^26^), among healthy adults without HIV, aged 18–50 years without chronic comorbidities, to test whether 12 weeks of NRTI exposure is associated with coordinated changes in DNA methylation–based measures of biological aging. We evaluated a comprehensive panel of first-, second-, and third-generation epigenetic clocks, organ-system–specific aging estimates, our retroelement-focused epigenetic clock^27^, DNA methylation proxies of inflammatory biomarkers^28^, and DNA methylation-based immune cell deconvolution^29^ to evaluate the breadth, specificity, and coherence of any treatment-associated epigenetic signal of two distinct NRTI regimens. Here, we demonstrate regimen-specific gerotherapeutic activity of NRTIs, serving as a proof-of-concept in humans and suggesting that intracellular drug exposure may be an important determinant of the biological aging response within this drug class.

## RESULTS

### Posthoc epigenetic analysis of two separate randomized, directly observed dosing pharmacokinetic NRTI regimen studies

We quantified biological age using established first-^30,31^, second-^32,33^, and third-generation epigenetic clocks^34,35^ using DNAm data generated from biobanked dried blood spots obtained from 79 total participants across two separate pharmacokinetic clinical trials (FTC/TAF n=36, FTC/TDF n=43) using epigenetic DNA-methylation (DNAm) data obtained at baseline (pre-drug) and after 12 weeks of continuous dosing (**Fig. 1**). We also evaluated versions of 1^st^ and 2^nd^ generation clocks built from DNAm principal components (PCs) (termed ‘PC clocks’) that show enhanced technical reliability and stability in longitudinal study designs such as randomized clinical trials^36^. Additionally, organ-specific aging estimates were derived for blood, brain, heart, hormone, immune, inflammation, kidney, liver, lung, metabolic, and musculoskeletal systems^37^. DNA methylation–based proxies of inflammatory biomarkers (DNAm CRP, DNAm HbA1c, and relative IL-6 levels) were also assessed^28^. The two separate clinical trials were broadly comparable in demographic and clinical characteristics, with no statistically significant differences in sex distribution (TAF: 50% male; TDF: 47% male; *p* = 0.934), weight (72.2 ± 14.7 vs. 80.7 ± 22.9 kg; *p* = 0.062), or age (28.4 ± 6.0 vs. 30.9 ± 7.3 years; *p* = 0.115) (**Table 1**).

**Figure 1.**
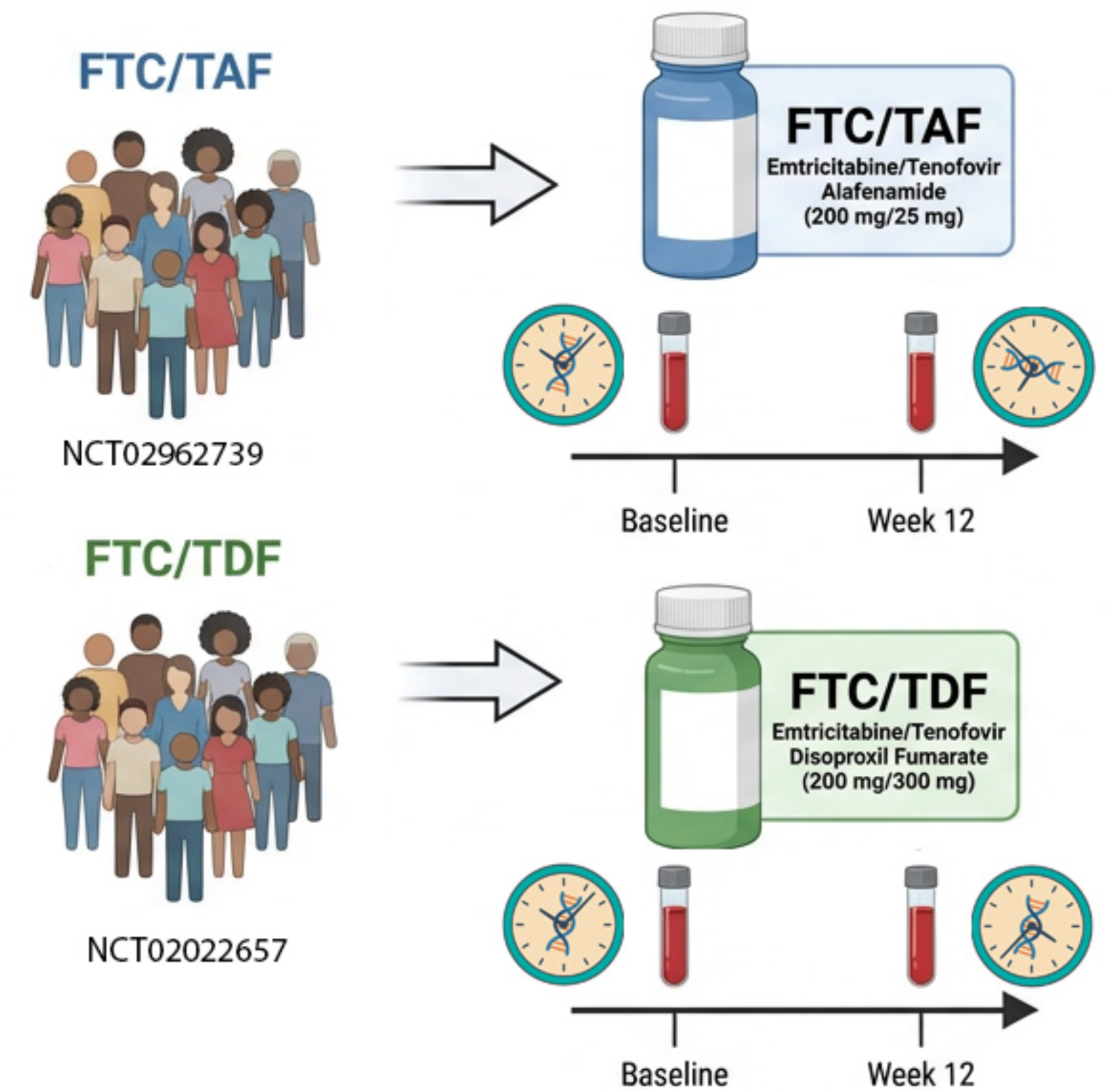
Study design overview. Post hoc epigenetic aging analyses were conducted using longitudinal specimens from two separate randomized, directly observed dosing pharmacokinetic studies: FTC/TAF (emtricitabine/tenofovir alafenamide, 200 mg/25 mg; NCT02962739) and FTC/TDF (emtricitabine/tenofovir disoproxil fumarate, 200 mg/300 mg; NCT02022657). Healthy adults without HIV aged 18–50 years were enrolled in each study. Dried blood spots were examined at baseline and week 12 for DNA methylation profiling and intracellular drug concentrations were measured at weekly or biweekly intervals.

**Table 1:** Baseline characteristics of the reverse transcriptase inhibitor clinical trial populations.

| Clinical Trial | TAF/FTC (N = 36) | TDF/FTC (N = 43) |
| --- | --- | --- |
| **Demographics** |  |  |
| Age (years) | 28.4 ± 6.0 | 30.9 ± 7.3 |
| Median [IQR] | 29 [24–32] | 29 [26–34] |
| Sex at birth, n (%) |  |  |
| Female | 18 (50%) | 23 (53%) |
| Male | 18 (50%) | 20 (47%) |
| Race, n (%) |  |  |
| White | 30 (83%) | 24 (56%) |
| Black or African-American | 5 (14%) | 7 (16%) |
| Other/Unknown/Multiracial | 1 (3%) | 12 (28%) |
| Hispanic or Latino, n (%) | 7 (19%) | 14 (33%) |
| **Anthropometrics** |  |  |
| Height (inches) | 67.2 ± 3.7 | 66.9 ± 4.2 |
| Weight (kg) | 72.2 ± 14.7 | 80.7 ± 22.9 |
| **Hematology** |  |  |
| Hematocrit (%) | 44.7 ± 3.4 | 42.8 ± 3.3 |
| Hemoglobin (g/dL) | 15.0 ± 1.3 | 14.3 ± 1.4 |
| Platelet count (×10<sup>9</sup>/L) | 259 ± 46 | 251 ± 57 |
| MCV (fL) | 89.4 ± 3.9 | 89.3 ± 5.0 |
| **Serum Chemistry** |  |  |
| Serum creatinine (mg/dL) | 0.8 ± 0.2 | 0.9 ± 0.2 |
| Potassium (mmol/L) | 3.9 ± 0.3 | 4.1 ± 0.3 |
| Glucose (mg/dL) | 84.9 ± 9.4 | 86.4 ± 13.7 |
| Bilirubin (mg/dL) | 0.7 ± 0.3 | 0.6 ± 0.4 |
| ALT (IU/L) | 21.8 ± 15.4 | 18.2 ± 8.4 |
| AST (IU/L) | 20.8 ± 8.0 | 18.5 ± 4.8 |

Values are mean ± SD unless otherwise indicated. IQR, interquartile range; MCV, mean corpuscular volume; ALT, alanine aminotransferase; AST, aspartate aminotransferase; TAF, tenofovir alafenamide; TDF, tenofovir disoproxil fumarate.

### FTC/TAF Slowed Epigenetic Aging Across Multiple DNA Methylation Clocks

Participants receiving FTC/TAF (N=36) demonstrated consistent reductions across the majority of epigenetic aging measures from baseline to week 12 (**Fig. 2**). Significant decreases were observed in DunedinPACE (−0.061, 95% CI: −0.11 to −0.01, p=0.019), indicating a measurable slowing of the pace of biological aging. Among epigenetic clock-based measures, the second-generation epigenetic clock PhenoAge showed the largest effect (−6.33 years, 95% CI: −10.91 to −1.74, p=0.008), followed by AdaptAge^35^ (−5.92 years, p=0.027), first-generation epigenetic clock Horvath^30^ (−4.27 years, p=0.023), PCHorvath2 (−3.81 years, p=0.020), PCHorvath1 (−3.63 years, p=0.015), and SystemsAge^37^ (−3.49 years, p=0.008). RetroClock^27^, which specifically captures the methylation state of retroelement loci including LINE-1s and HERVs, showed a trend toward reduction (−2.78 years, p=0.051). Additional epigenetic clocks evaluated including PCGrimAge (−1.39 years, p=0.069), PCHannum (−2.74 years, p=0.081), Hannum (−2.37 years, p=0.119), and PCPhenoAge (−2.49 years, p=0.145) showed directionally consistent reductions. We did not observe any notable differences in DamAge (+2.10 years, p=0.252), OMICmAge (+0.35 years, p=0.638), and IntrinsicCapacity clock (−0.56 years, p=0.577). Collectively, our findings demonstrate a significant, coordinated reduction in DNAm-based epigenetic aging with FTC/TAF.

**Figure 2.**
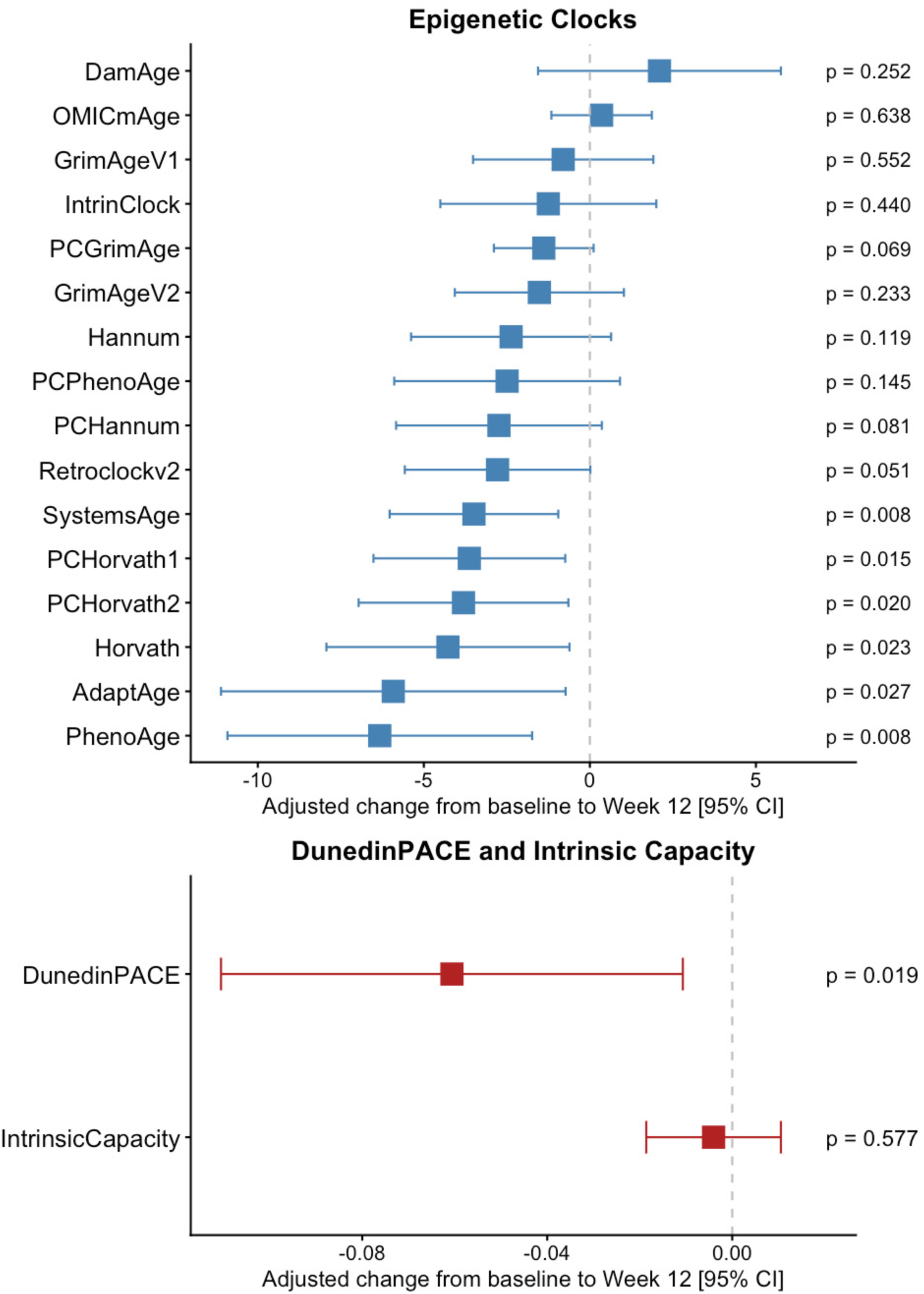
FTC/TAF is associated with broad deceleration of DNA methylation–based aging measures. Forest plots show adjusted within-participant change from baseline to Week 12 (Week 12 − baseline) for DNA methylation–derived biomarkers of aging in the FTC/TAF arm. Point estimates (squares) and 95% confidence intervals (horizontal bars) are from ANCOVA-style linear mixed-effects models including timepoint, age, and sex as fixed effects and a participant-level random intercept; the vertical dashed line denotes no change. Effects across 16 epigenetic clocks and on DunedinPACE (pace of aging) and the Intrinsic Capacity clock, shown on their native scales. Negative values indicate lower epigenetic age estimates or a slower pace of aging.

### FTC/TAF Reduced Epigenetic Proxies of Inflammation

To evaluate whether FTC/TAF modulated systemic inflammation, we quantified DNA methylation–derived proxies of circulating inflammatory and metabolic biomarkers from whole-blood methylation profiles. Specifically, we tested longitudinal change in the DNAm IL-6 score^38^ and GrimAgeV2-derived^28^ DNAm logCRP (with DNAm logA1C as a complementary metabolic/inflammation-adjacent marker) as prespecified readouts of inflammation-related biology, and compared within-participant shifts over follow-up in the FTC/TAF group. Consistent with the hypothesis that NRTIs may attenuate age-associated systemic inflammation^39–41^, DNA methylation–based proxies of inflammatory biomarkers showed concordant reductions in the FTC/TAF group (**Fig. 3**). DNAm-derived IL-6^38^ levels decreased significantly (−0.058 arbitrary unit (a.u.), 95% CI: −0.11 to −0.006, p=0.029), and DNAm C-reactive protein showed a trend toward reduction (−0.231 a.u., p=0.059). DNAm HbA1c also trended downward (−0.007 a.u., p=0.132). Together, these coordinated downward shifts in inflammation-related DNAm proxies are consistent with an anti-inflammatory signal during FTC/TAF exposure, supporting the proposed link between reverse transcriptase inhibition and reduced systemic inflammatory tone.

**Figure 3.**
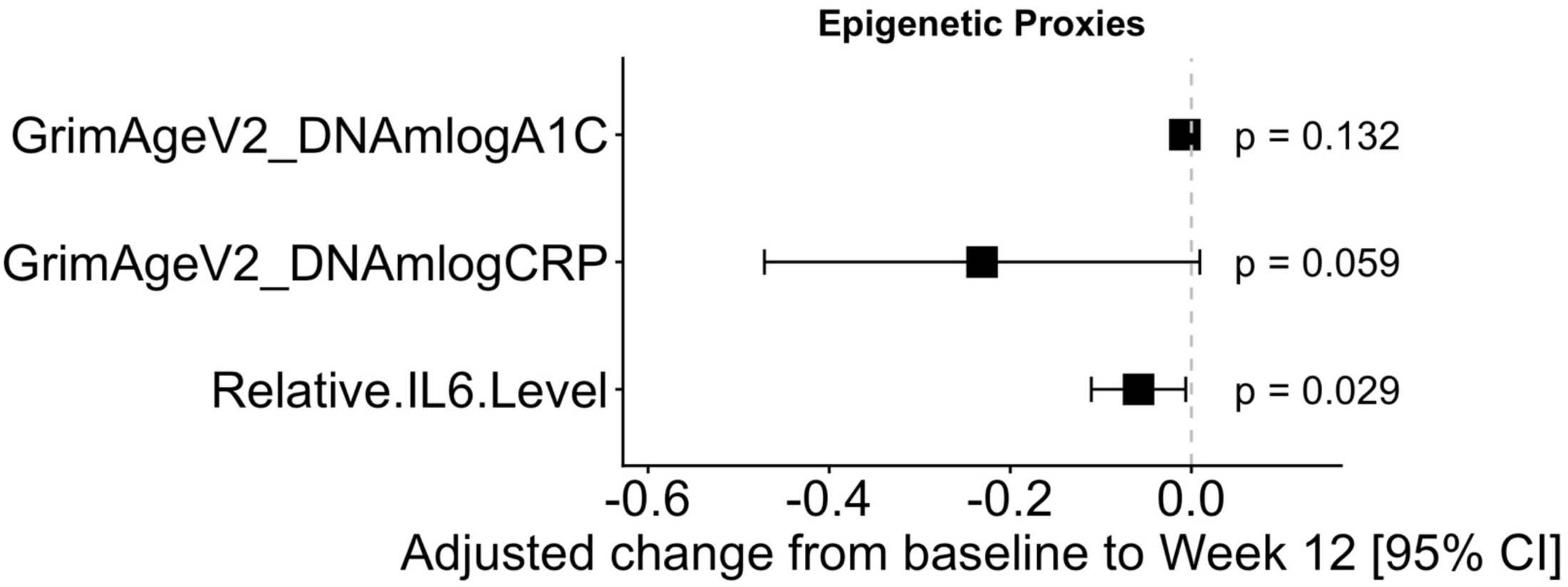
FTC/TAF is associated with reduced DNA methylation–based proxies of inflammatory and metabolic biomarkers. Forest plot shows adjusted within-participant change from baseline to week 12 (week 12 − baseline) in DNA methylation–derived proxy measures for HbA1c (DNAm logA1C), C-reactive protein (DNAm logCRP), and relative IL-6 levels. Squares denote model-estimated mean differences and horizontal bars indicate 95% confidence intervals from ANCOVA-style linear mixed-effects models adjusting for age and sex (with participant-level random intercepts). The vertical dashed line indicates no change; negative values indicate lower inferred biomarker levels. FTC/TAF was associated with a significant reduction in the IL-6 proxy and a trend toward reduced DNAm CRP, with no significant change in the DNAm HbA1c proxy.

### FTC/TAF Conferred Multi-Organ Deceleration of System-Level Epigenetic Aging

To determine whether the epigenetic aging signal under FTC/TAF was localized or distributed across physiological domains, we applied the Systems Age framework, which decomposes whole-blood DNA methylation into 11 clinically anchored system-age estimates (heart, lung, kidney, liver, brain, immune, inflammatory, blood, musculoskeletal, hormone and metabolic) derived using clinical biomarkers, functional assessments and mortality risk^37^. This allowed us to test whether FTC/TAF was associated with coordinated epigenetic aging shifts across multiple physiological systems. Across the 12-week FTC/TAF exposure period, system-specific epigenetic clocks exhibited broad, multi-domain decreases, estimated using ANCOVA-style linear mixed-effects models with age and sex as covariates and participant-level random intercepts (**Fig. 4**). Systems derived blood age showed the most robust decrease (−3.44 years, p<0.001), followed by Systems metabolic age (−3.38 years, p=0.005), Systems heart age (−3.23 years, p=0.009), Systems hormone age (−3.05 years, p=0.033), Systems inflammation age (−2.42 years, p=0.015), and Systems brain age (−2.18 years, p=0.039). Systems kidney age (−1.98 years, p=0.147), systems musculoskeletal age (−1.28 years, p=0.177), systems liver age (−1.14 years, p=0.317), and systems lung age (−0.78 years, p=0.484) showed directionally consistent but non-significant reductions. These results indicate that FTC/TAF was associated with a distributed, multisystem shift toward younger Systems Age estimates over 12 weeks.

**Figure 4.**
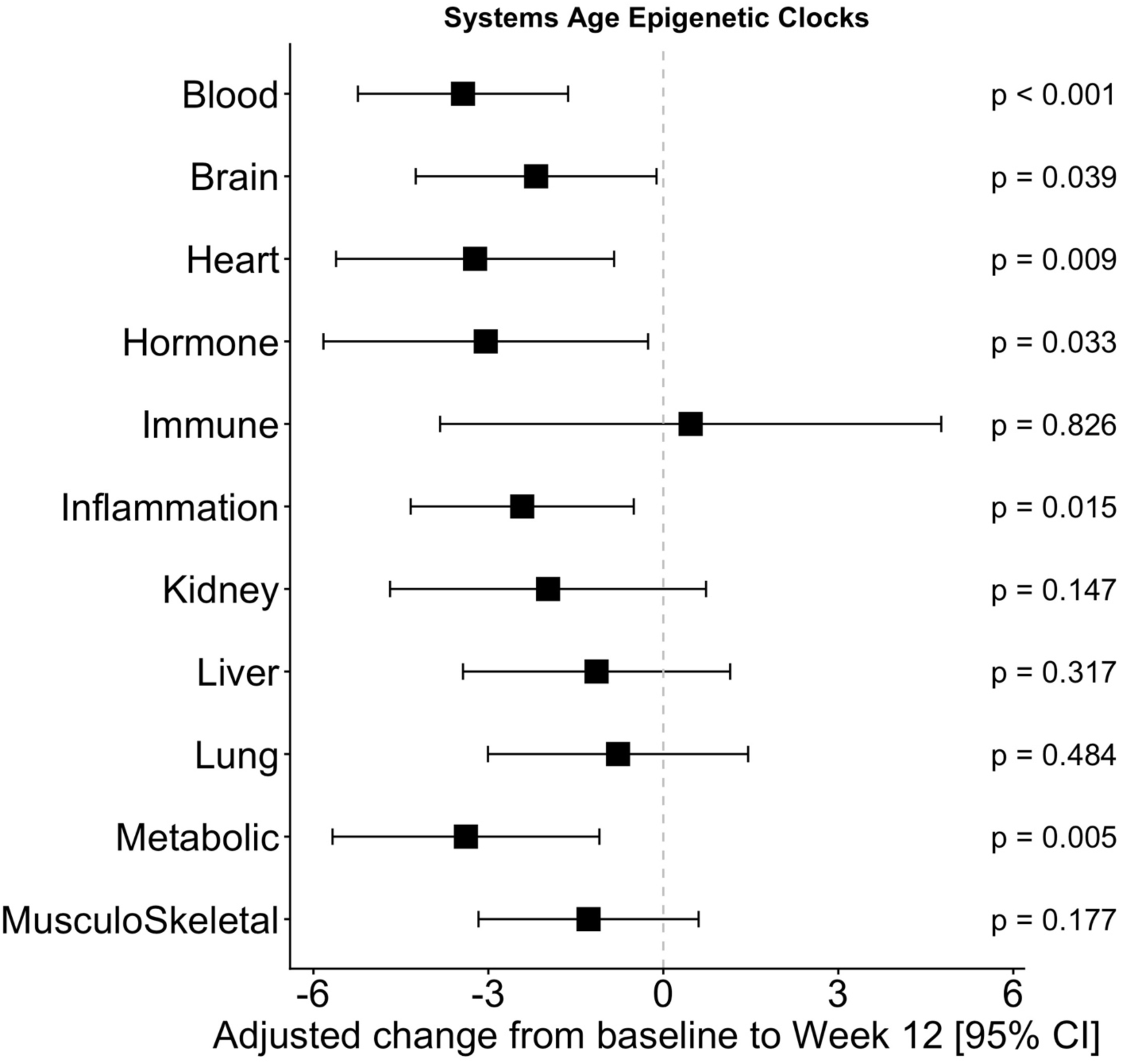
FTC/TAF is associated with deceleration of system-level epigenetic aging across multiple systems clocks. Forest plot shows adjusted within-participant change from baseline to week 12 (week 12 − baseline) in DNA methylation–derived system-specific epigenetic age estimates, including blood, brain, heart, hormone, immune, inflammation, kidney, liver, lung, metabolic, and musculoskeletal clocks. Squares denote model-estimated mean differences and horizontal bars indicate 95% confidence intervals from ANCOVA-style linear mixed-effects models adjusting for age and sex (with participant-level random intercepts). The vertical dashed line indicates no change; negative values indicate lower system-level epigenetic age (slower aging).

### FTC/TAF Modulated Immune Cell Composition

We next assessed whether FTC/TAF was accompanied by shifts in immune cell composition from whole-blood DNA methylation profiles using an immune cell type reference-based deconvolution approach^29^. DNA methylation–based immune cell deconvolution in the FTC/TAF group revealed significant shifts in the inferred proportions of specific immune cell types (**Fig. 5**). We observed that the proportion of naive CD4+ T cells (CD4Tnv) significantly increased over 12 weeks (+0.050 percent, p=0.008), consistent with a shift toward a more naïve T cell immune compartment. Regulatory T cells (Tregs) also significantly increased over 12 weeks (+0.016 percent, p=0.049), while neutrophils significantly decreased (−0.066 percent, p=0.048), indicating a shift toward a more immunoregulatory and less granulocyte-weighted immune-cell composition following FTC/TAF. Other cell populations including basophils, memory B cells, naive B cells, memory CD4+ T cells, memory and naive CD8+ T cells, eosinophils, monocytes, and NK cells did not change significantly. The increase in naive CD4+ T cells is particularly notable given that the decline in naive T cell output is a hallmark of immune aging, and interventions that restore naive T cell pools have been proposed as markers of successful geroprotective strategies^42^.

**Figure 5.**
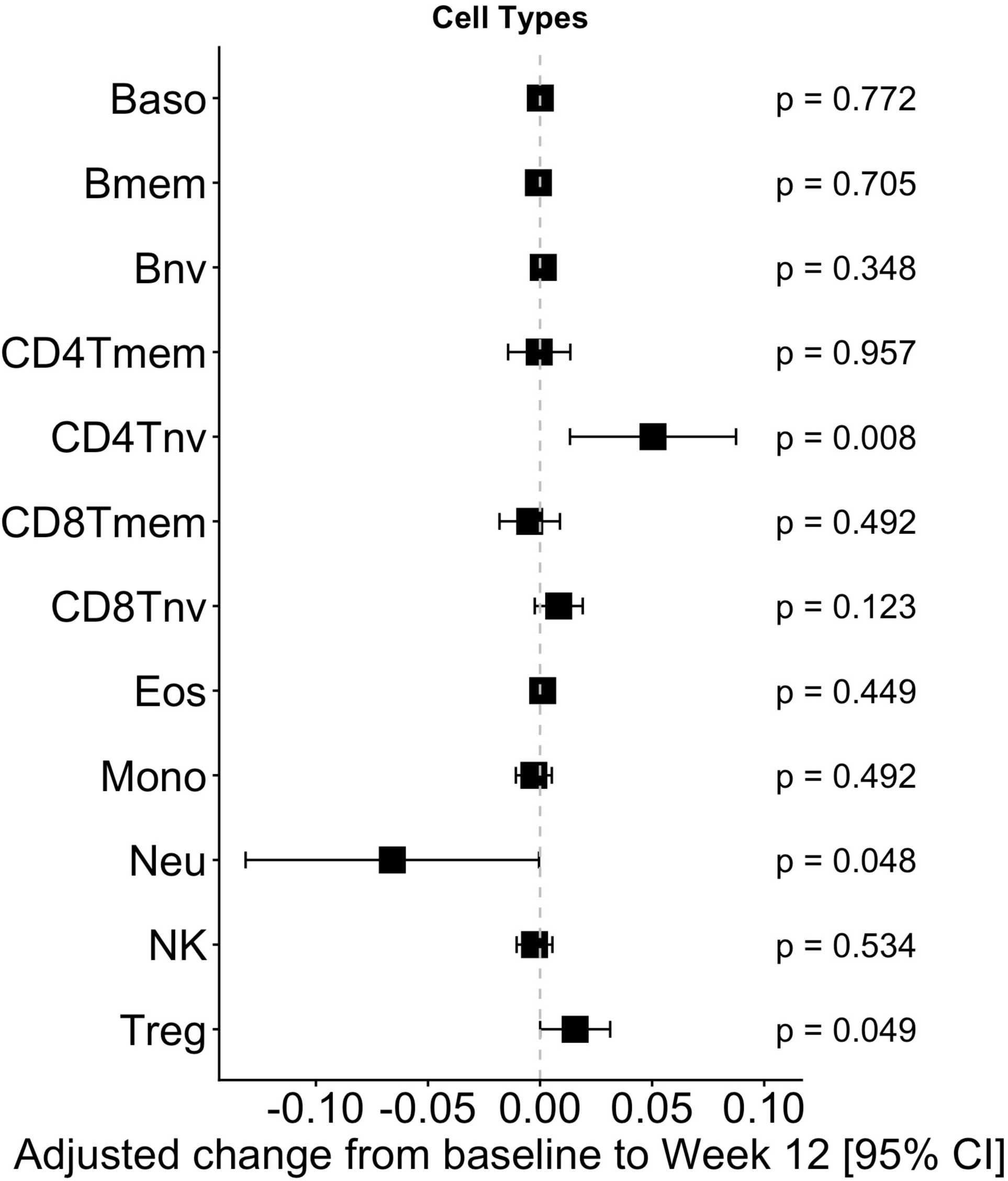
FTC/TAF is associated with shifts in DNA methylation–inferred immune cell composition. Forest plot shows adjusted within-participant change from baseline to week 12 (week 12 − baseline) in estimated immune cell proportions inferred from whole-blood DNA methylation profiles. Squares denote model-estimated mean differences and horizontal bars indicate 95% confidence intervals from ANCOVA-style linear mixed-effects models adjusting for age and sex (with participant-level random intercepts). The vertical dashed line indicates no change; positive values indicate increased inferred proportion and negative values indicate decreased inferred proportion. FTC/TAF was associated with increased naïve CD4 T cells (CD4Tnv) and regulatory T cells (Treg), alongside reduced neutrophils (Neu), with other cell subsets showing no significant change.

### FTC/TDF Did Not Change Epigenetic Aging Across Multiple DNA Methylation Clocks

To test whether the multi-clock epigenetic age reductions observed with 12 weeks of FTC/TAF in healthy people without HIV reflect a broader NRTI-associated signal or instead may depend on regimen-specific pharmacology, we applied the same analytic pipeline to a separate randomized, directly observed pharmacokinetic study of 43 participants with comparable population characteristics and 12-week follow-up. In contrast to findings from our FTC/TAF study, participants in the FTC/TDF trial exhibited a markedly different epigenetic aging profile over 12 weeks. Across the 12-week FTC/TDF exposure period, epigenetic aging measures showed no evidence of a coordinated deceleration or reduction (**Fig. 6**). We observed that OMICmAge showed a significant positive shift (+0.63, p=0.014), PCPhenoAge trended upward (+0.77, p=0.099), and DunedinPACE trended higher (+0.024, p=0.063). Notably, we observed that the IntrinsicCapacity epigenetic clock estimate significantly decreased (−0.005, p=0.019), diverging in direction from all other epigenetic clocks observed as unchanged or shifted modestly upward. Moreover, epigenetic inflammatory proxies in the FTC/TDF study showed no significant changes: DNAm CRP (−0.044, p=0.435), DNAm HbA1c (+0.006, p=0.220), and IL-6 (+0.014, p=0.193) were all non-significant (**Fig. 7**). Furthermore, Systems Age epigenetic clocks under FTC/TDF did not show any significant change over 12 weeks (**Fig. 8**). Immune cell deconvolution in the FTC/TDF group showed a distinct pattern, with a significant increase in the proportion of DNA-methylation inferred neutrophils (+0.042, p=0.039) and a significant decrease in basophils (−0.002, p=0.041) (**Fig. 9**). Other cell types showed no significant changes following 12 weeks of FTC/TDF.

**Figure 6.**
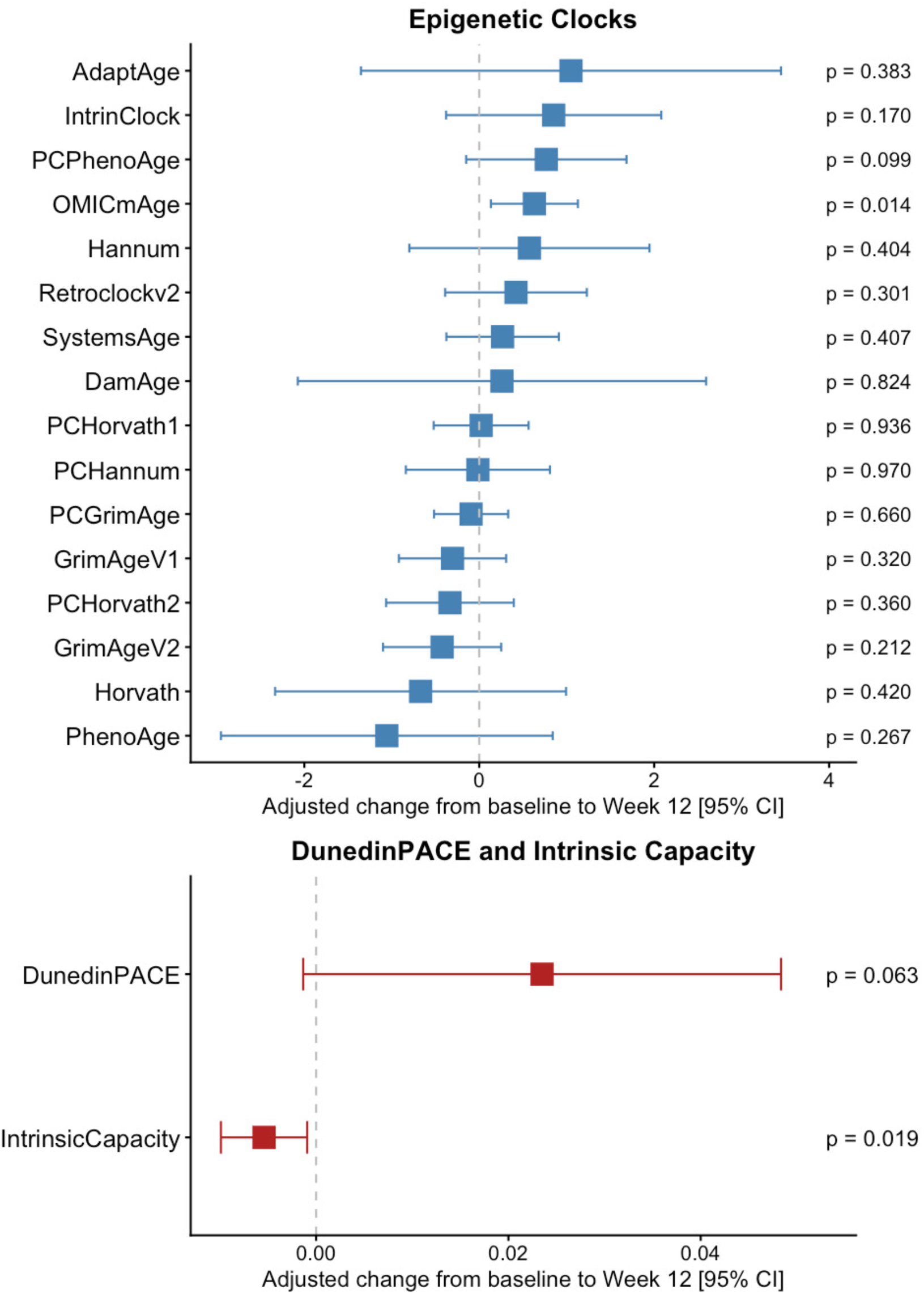
FTC/TDF shows minimal and inconsistent effects on DNA methylation–based aging measures. Forest plots show adjusted within-participant change from baseline to Week 12 (Week 12 − baseline) for DNA methylation–derived biomarkers of aging in the FTC/TDF arm. Point estimates (squares) and 95% confidence intervals (horizontal bars) are from ANCOVA-style linear mixed-effects models including timepoint, age, and sex as fixed effects and a participant-level random intercept; the vertical dashed line denotes no change. Effects across 16 epigenetic clocks and on DunedinPACE (pace of aging) and the Intrinsic Capacity clock, shown on their native scales. Negative values indicate lower epigenetic age estimates or a slower pace of aging.

**Figure 7.**
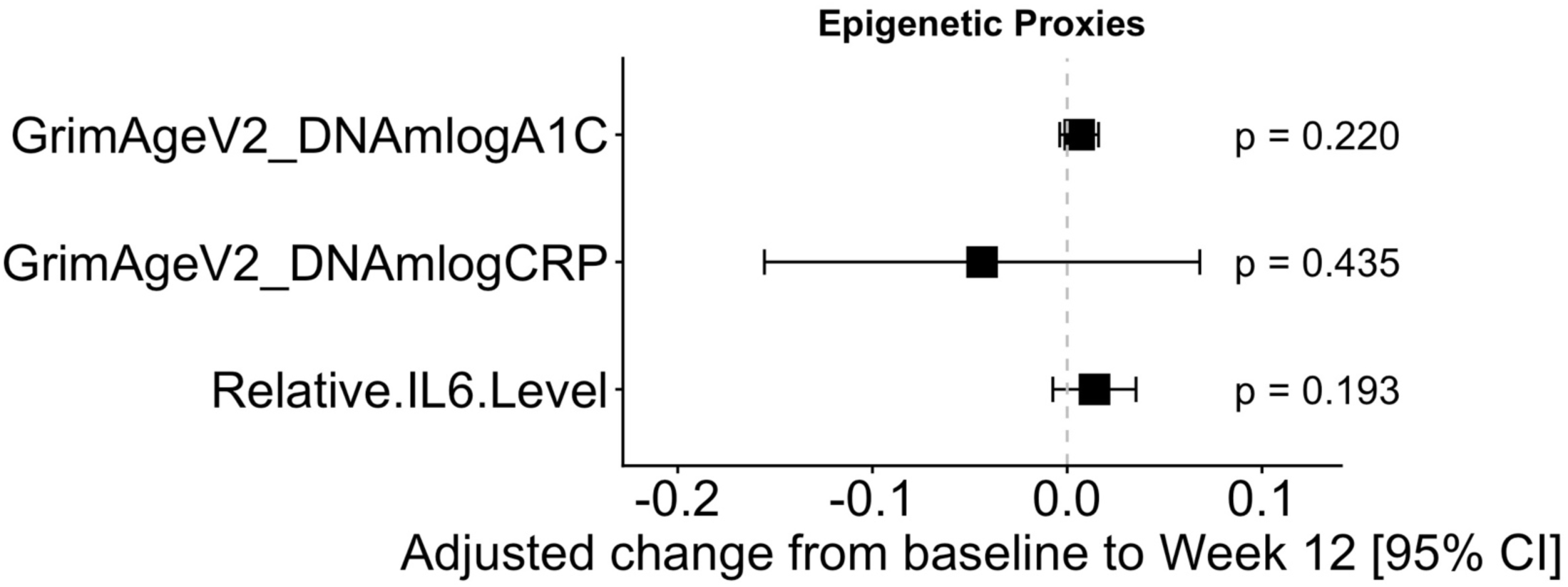
FTC/TDF does not measurably alter DNA methylation–based proxies of inflammatory and metabolic biomarkers. Forest plot shows adjusted within-participant change from baseline to week 12 (week 12 − baseline) in DNA methylation–derived proxy measures for HbA1c (DNAm logA1C), C-reactive protein (DNAm logCRP), and relative IL-6 levels in the FTC/TDF arm. Squares denote model-estimated mean differences and horizontal bars indicate 95% confidence intervals from ANCOVA-style linear mixed-effects models adjusting for age and sex (with participant-level random intercepts). The vertical dashed line indicates no change; negative values indicate lower inferred biomarker levels. All proxy estimates were centered near zero with non-significant changes across biomarkers.

**Figure 8.**
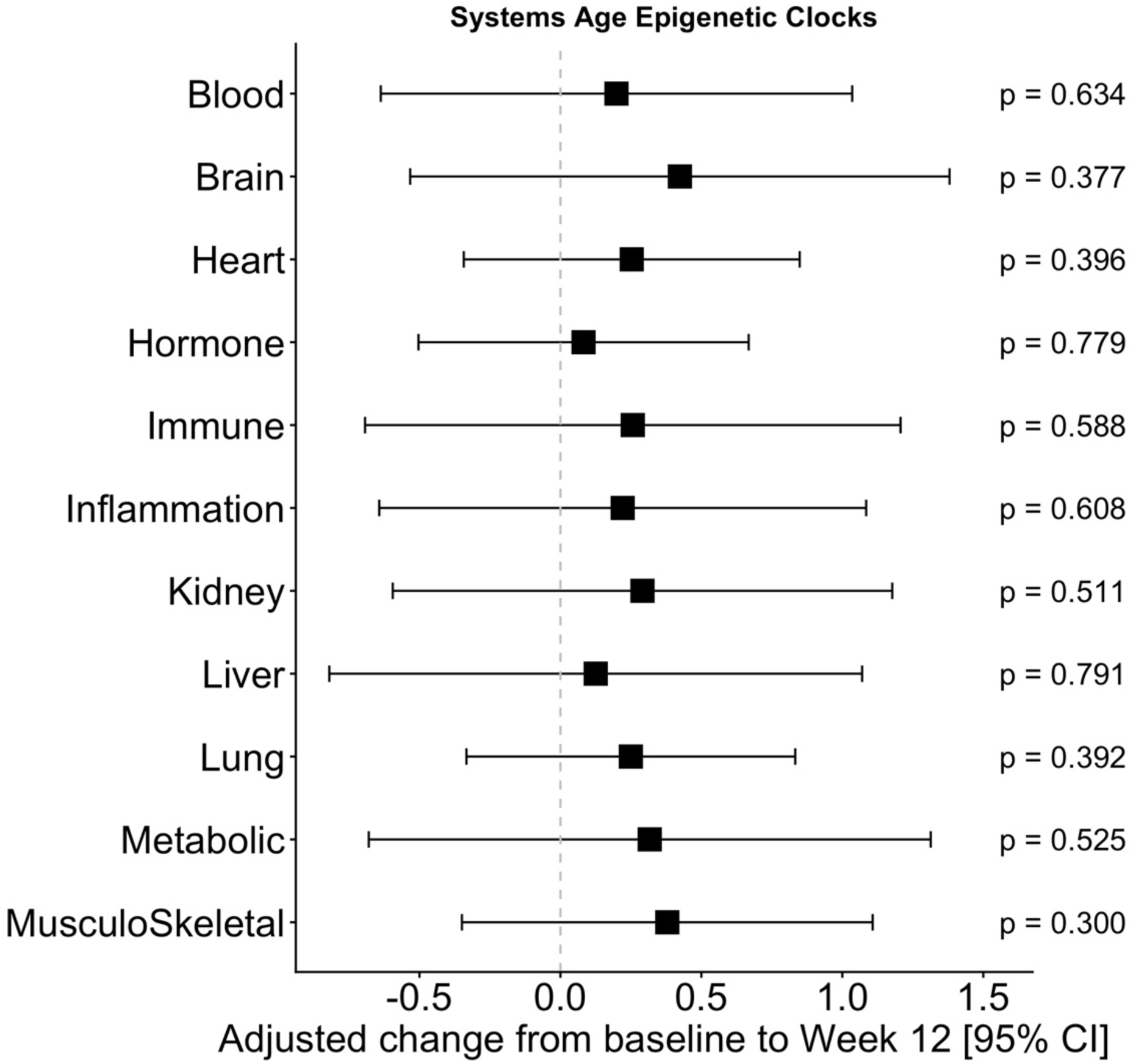
FTC/TDF does not measurably alter system-level epigenetic aging across systems clocks. Forest plot shows adjusted within-participant change from baseline to Week 12 (Week 12− baseline) in DNA methylation–derived system-specific epigenetic age estimates, including blood, brain, heart, hormone, immune, inflammation, kidney, liver, lung, metabolic, and musculoskeletal clocks. Squares denote model-estimated mean differences and horizontal bars indicate 95% confidence intervals from ANCOVA-style linear mixed-effects models adjusting for age and sex (with participant-level random intercepts). The vertical dashed line indicates no change; negative values indicate lower system-level epigenetic age (slower aging). Estimates were centered near zero across systems, with no significant multi-organ deceleration observed.

**Figure 9.**
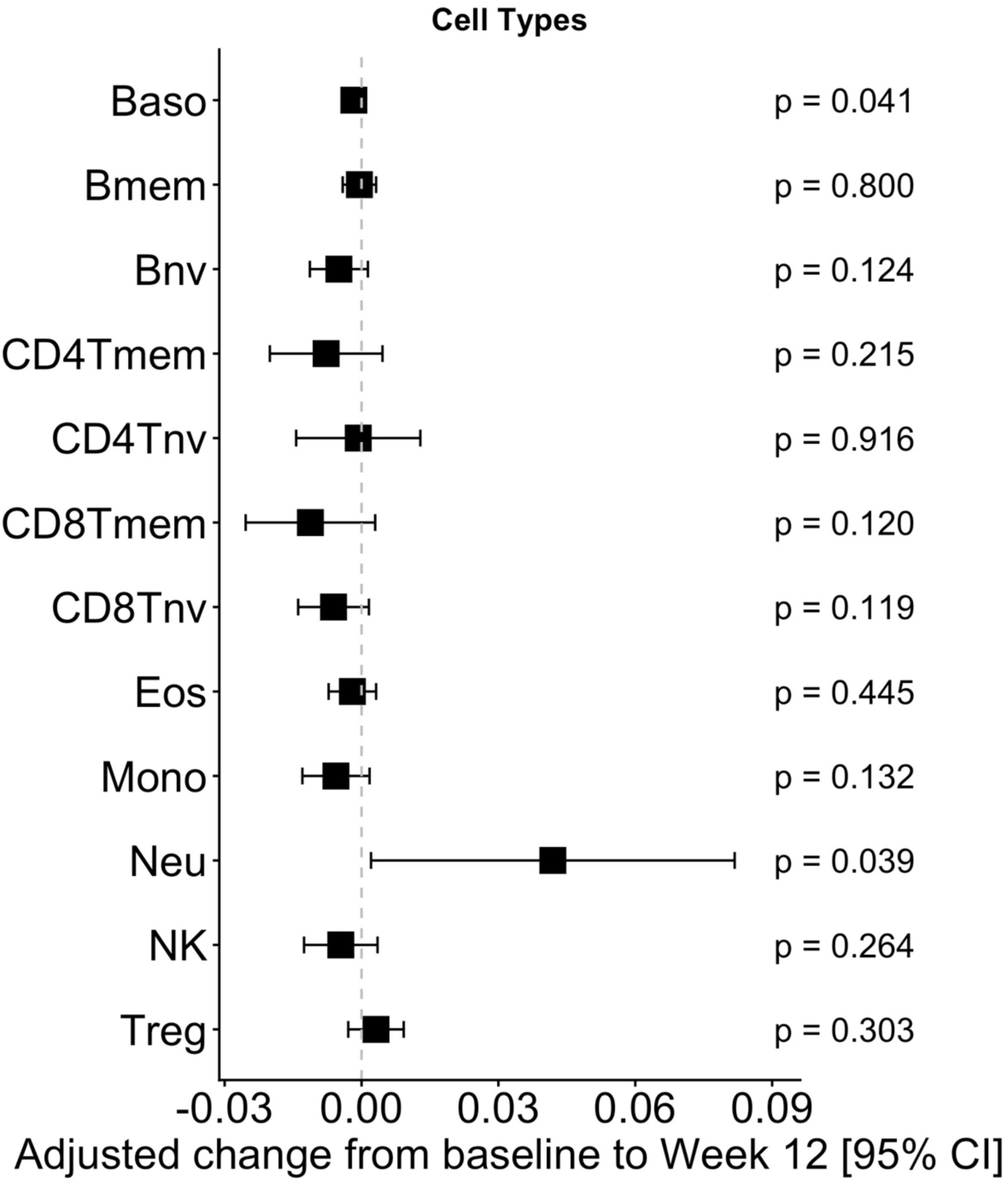
FTC/TDF is associated with modest shifts in DNA methylation–inferred immune cell composition. Forest plot shows adjusted within-participant change from baseline to week 12 (week 12 − baseline) in estimated immune cell proportions inferred from whole-blood DNA methylation profiles in the FTC/TDF arm. Squares denote model-estimated mean differences and horizontal bars indicate 95% confidence intervals from ANCOVA-style linear mixed-effects models adjusting for age and sex (with participant-level random intercepts). The vertical dashed line indicates no change; positive values indicate increased inferred proportion and negative values indicate decreased inferred proportion. FTC/TDF was associated with an increase in neutrophils and a decrease in basophils, while other inferred cell subsets showed no significant change.

**Figure 10.**
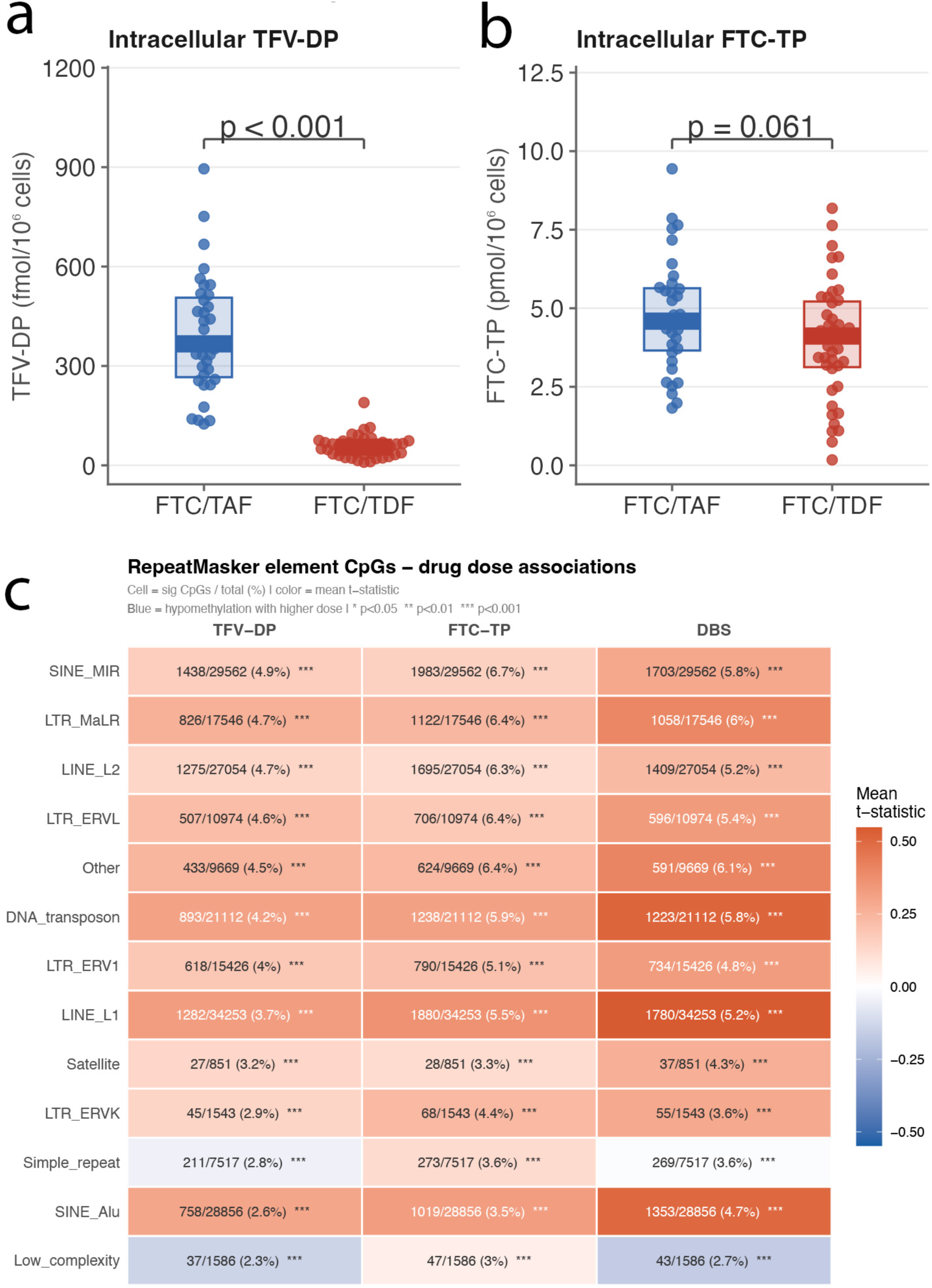
Regimen-specific intracellular metabolite exposure and dose-associated DNA methylation changes at RepeatMasker CpGs. **A.** Intracellular tenofovir diphosphate (TFV-DP) concentrations in participants receiving FTC/TAF or FTC/TDF. Points represent individual participants; center lines indicate the median; boxes indicate the interquartile range (IQR); and whiskers indicate 1.5 × IQR. TFV-DP concentrations were substantially higher in the FTC/TAF group than in the FTC/TDF group, consistent with the known pharmacokinetic separation between the two tenofovir prodrugs. **B.** Intracellular emtricitabine triphosphate (FTC-TP) concentrations in participants receiving FTC/TAF or FTC/TDF. FTC-TP levels were similar between groups, consistent with the shared 200mg emtricitabine backbone. **C.** In the FTC/TAF arm, heat map summarizing associations between drug dose biomarkers and DNA methylation at CpGs annotated to RepeatMasker element classes. Columns correspond to TFV-DP, FTC-TP and DBS, and rows to repeat classes. Numbers within cells indicate the number of significant CpGs relative to the total number tested for each class, with percentages in parentheses. Cell colour denotes the mean *t*-statistic across significant CpGs, with warmer colours indicating more positive associations and cooler colours indicating more negative associations; blue indicates hypomethylation with higher drug dose. Across multiple repeat classes, including SINE, LINE, LTR, satellite and DNA transposon elements, higher intracellular drug exposure was associated with widespread shifts in DNA methylation levels, supporting a dose-responsive pharmacoepigenetic effect at repeat-associated CpGs. Asterisks denote significance (*P* < 0.05, \**P* < 0.01, \*\**P* < 0.001).

### Regimen-specific gerotherapeutic activity is consistent with divergent intracellular tenofovir-diphosphate exposure

We next asked whether interindividual variation in intracellular drug exposure was associated with DNA methylation change over the 12-week period at CpGs mapping to repetitive elements. Within the subset of participants who underwent paired DNA methylation profiling for epigenetic age analysis, intracellular TFV-DP concentrations were substantially higher in FTC/TAF-treated participants than in FTC/TDF-treated participants (**Fig. 9A**), whereas FTC-TP concentrations were similar between regimens (**Fig. 9B**), indicating that intracellular tenofovir loading was the principal pharmacokinetic feature distinguishing the two arms. Within the FTC/TAF study, using a paired delta-EWAS framework, we modeled post-minus-pre M-value change at each CpG as a function of standardized post-dose TFV-DP, FTC-TP, or dried blood spot (DBS) exposure and observed broad concentratoin-associated methylation shifts across RepeatMasker-annotated loci. The level of TFV-DP exposure in cells was associated with significant DNA methylation change over the 12-week period at 8,350 CpGs (**Fig. 9C**). Within individual repeat classes, these signals involved 2.3–6.7% of CpGs from the DNA methylation array that spanned SINE, LINE, LTR, DNA transposon and satellite elements. Most repeat classes exhibited positive mean *t*-statistics, consistent with higher TFV-DP exposure being associated with higher methylation levels at repeat-associated CpGs. Because DNA methylation is a core epigenetic feature of transposable element repression, these findings raise the possibility that greater intracellular TFV-DP exposure may promote or stabilize cellular states that favor transposable element silencing.

## DISCUSSION

In this post hoc longitudinal analysis of two randomized, directly observed dosing pharmacokinetic studies, 12 weeks of the FDA-approved NRTI regimen FTC/TAF was associated with coordinated decreases across multiple DNA methylation–based measures of biological aging in healthy adults without HIV or chronic comorbidities. The signal was not confined to a single clock family: we observed reductions spanning first-generation chronological age predictors, second-generation morbidity- and mortality-trained clocks, and the pace-of-aging measure DunedinPACE, with concordant shifts across several system-level aging estimates. In parallel, DNA methylation–based proxies of inflammatory tone (notably epigenetic IL-6, with a trend for DNAm CRP) declined, and immune deconvolution suggested a shift toward a more “youthful” circulating profile, including increased naïve CD4 T cells and reduced neutrophils. Together, these results suggest that 12 week FTC/TAF exposure is associated with coordinated shifts in reducing/slowing multiple DNA methylation–based aging measures, and that intracellular exposure to the active form of tenofovir (tenofovir-diphosphate) may be a relevant explanatory factor.

The divergent effects of these two NRTI regimens are noteworthy in the context of geroscience repurposing efforts. FTC/TAF and FTC/TDF share emtricitabine (a close analog to 3TC (lamivudine)), but differ in the tenofovir prodrug, producing markedly different exposure profiles: the TAF prodrug is preferentially metabolized in liver (by carboxylesterase-1) and immune tissues (by cathepsin-A) thereby generating high intracellular tenofovir-diphosphate in these compartments with lower systemic tenofovir exposure^43,44^, whereas the TDF prodrug is metabolized upon absorption (by carboxylesterase-2), first pass (carboxylesterases-1 and 2), and plasma esterases yielding high plasma tenofovir, which is associated with off-target renal and bone effects, with comparatively lower intracellular tenofovir-diphosphate concentrations in immune tissues. For example, FTC/TAF (200mg/25mg) produces approximately 7-fold higher active tenofovir-diphosphate in PBMC compared with FTC/TDF (200mg/300mg) concurrently with 1/10^th^ the concentration of circulating tenofovir in plasma^23^. Because (i) peripheral blood immune cells are the substrate for DNA methylation profiling and (ii) these same cells are a major arena in which retrotransposon de-repression can generate immunostimulatory nucleic acids, the intracellular TFV-DP enrichment achieved by TAF in these tissues provides a plausible exposure–response rationale for the stronger epigenetic aging signal. Importantly, our retrospective design does not permit a causal, head-to-head comparison between trials, but the qualitative contrast and broad, directionally consistent deceleration under FTC/TAF versus minimal changes under FTC/TDF given similar participant populations supports the idea that immune cell targeting of tenofovir-diphosphate with TAF may be a key determinant of any geroprotective effect for this drug class.

These findings support and extend a retrotransposon-centric model of aging in which progressive erosion of chromatin architecture and epigenetic repression permits increased expression of LINE-1 and endogenous retroviruses, generating nucleic acid species that engage innate immune sensing pathways, including cGAS–STING and RIG-I/MDA5–MAVS, and thereby reinforce type I interferon signaling and senescence-associated inflammatory programs. In this context, our study provides translational human evidence that FTC/TAF may modulate this axis: beyond the observed slowing across multiple epigenetic aging measures, short-term treatment was also associated with reductions in epigenetic proxies of inflammatory biomarkers, suggesting an anti-inflammatory effect independent of clock-derived outcomes. The directionally consistent effect on RetroClock^27^, the first retroelement-focused epigenetic clock, further raises the possibility that FTC/TAF may influence the epigenetic state of transposable element loci themselves, although this signal requires replication and may have been attenuated by the young age of the cohort and the short 12-week exposure window, both of which likely limit the dynamic range of transposable element activity. At the same time, methylation-based immune cell-type deconvolution^29^ indicated shifts toward a more youthful immune profile, including increased naïve CD4+ T cells and regulatory T cells and reduced neutrophils, changes that may both reflect and contribute to the broader epigenetic clock deceleration. Because whole-blood methylation signatures are sensitive to cell-mixture effects^45^, future studies should distinguish cell-intrinsic methylation remodeling from compositional immune changes using measured hematologic phenotyping, cell-type-specific immune profiling, or single-cell epigenomic approaches, while also testing longer FTC/TAF exposure and direct molecular readouts of transposable element activity in older or higher-risk populations, where baseline derepression may be greater and treatment effects more readily detectable.

These findings have direct implications for ongoing efforts to develop novel reverse transcriptase inhibitors as gerotherapeutics. Our data suggest that intracellular tenofovir-diphosphate concentration in immune cells not systemic RT inhibition per se is the critical pharmacological determinant of epigenetic aging benefit. Candidate gerotherapeutics targeting retrotransposon reverse transcription should therefore demonstrate favorable intracellular pharmacokinetics in immune cell compartments comparable to or exceeding those achieved by FTC/TAF before advancing to costly clinical development. More broadly, these data arrive at a pivotal moment for translational geroscience, as the field shifts from descriptive aging biology to intervention-based healthspan testing. ARPA-H’s PROactive Solutions for Prolonging Resilience (PROSPR) program includes a retrotransposon-focused reverse transcriptase strategy using a formerly developed HIV NRTI being repurposed for transposon biology^46^. In parallel, XPRIZE Healthspan is a global competition challenging teams to deliver, within 1 year, single or combinations of therapeutics that restore muscle, cognitive and immune function by at least 10 years in older adults^47^. In this emerging clinical trial landscape, clinical pharmacology may be a key translational determinant of success for retrotransposon-targeted gerotherapeutics.

Several limitations warrant consideration. This was a post hoc analysis of two independent pharmacokinetic studies that were neither designed nor powered for aging end points, with modest sample sizes and only 12 weeks of follow-up. There was no placebo group, and because FTC/TAF and FTC/TDF were studied separately, the data do not support a formal head-to-head comparison between regimens. As part of the original study designs for the clinical trials, a subset of the participants received intermittent dosing schedules equivalent to one third or two-thirds of standard daily dosing, so it remains possible that the biological effects observed here were attenuated by lower dose exposure. However, aspects of the design strengthen the interpretability of the findings: the cohorts were young and otherwise healthy, limiting confounding by chronic disease, HIV infection and polypharmacy; drug administration was directly observed; and participant-level intracellular pharmacology data were available, allowing DNA methylation changes to be interpreted in the context of measured exposure. Another limitation is that multiple epigenetic aging outcomes were examined without formal correction for multiple testing, and the results should therefore be viewed as exploratory. DNA methylation was profiled from whole blood collected as dried blood spots. Although this approach cannot resolve tissue-specific effects, it is minimally invasive, well suited to repeated sampling and highly scalable, which is an important practical advantage for future larger and more geographically diverse studies. Finally, because the study population was young and healthy, with a limited baseline burden of age-related molecular dysfunction, the size, persistence and generalizability of these effects in older populations or in those with greater retroelement derepression remain unknown.

Despite these limitations, our findings provide the first clinical evidence to our knowledge that an FDA-approved NRTI regimen is associated with slowing/reducing epigenetic measures of biological aging in healthy adults. Our results indicate that FTC/TAF showed consistent reductions across multiple independent epigenetic clock estimates and concordant decreases in epigenetic proxies of inflammation, whereas FTC/TDF did not. Also noteworthy is the directly-observed dosing study designs that assured adherence and measured exposures to the NRTI regimens over the 12 weeks of observation. These results support the emerging concept that pharmacological targeting of retrotransposon activity represents a viable gerotherapeutic strategy^12^ and provide rationale for prospective, randomized, placebo-controlled trials designed to evaluate NRTIs as interventions for biological aging in the general population. Such trials should integrate measures of drug exposure and adherence, pharmacokinetic/pharmacodynamic modeling of intracellular drug concentrations, direct transposable element readouts (L1 mRNA, ORF1p protein, retroelement-derived cytoplasmic DNA), and prespecified biological aging endpoints to rigorously test causality and generalizability. As the field of geroscience moves toward identifying clinically actionable interventions and seeks to repurpose FDA approved drugs into aging therapies, the well-characterized safety profile and global availability of licensed NRTI formulations position FTC/TAF as a compelling, immediately testable candidate for repurposing to slow biological aging and extend healthspan.

## Materials and Methods

### Study Design and Participants

Biospecimens were obtained from two separate randomized, crossover pharmacokinetic studies that enrolled healthy adults aged 18–50 years without HIV infection. Participants were required to have an estimated glomerular filtration rate ≥60 mL/min/1.73 m² (Modified Diet in Renal Disease equation), no history of nontraumatic bone fractures, and no medical conditions known to alter red blood cell kinetics (e.g., hemoglobinopathies, hemolysis). Because participants received less than daily dosing during portions of the study, individuals at high risk of HIV acquisition were excluded. Additional exclusion criteria included confirmed HIV or hepatitis B virus infection, pregnancy, urine protein ≥2+, and any condition that, in the opinion of the investigators, might interfere with study procedures or participant safety. The FTC/TDF study was conducted at the San Francisco Department of Health and University of Colorado Anschutz Medical Campus, approved by the respective Institutional Review Boards, and was registered on ClinicalTrials.gov (NCT02022657); specimens were biobanked under a Colorado Multiple Institutional Review Board approved protocol. The FTC/TAF study was conducted at the University of Colorado Anschutz Medical Campus under a Colorado Multiple Institutional Review Board approved protocol and was registered on ClinicalTrials.gov (NCT02962739). All participants provided written informed consent prior to enrollment including consent for using stored samples for future genetic and other testing.

Participants were randomized to two of three directly observed dosing regimens 33%, 67%, or 100% of daily dosing each sustained for 12 weeks and separated by a 12-week washout period, for a total study duration of approximately 36 weeks. The FTC/TAF study participants received emtricitabine 200 mg/tenofovir alafenamide 25 mg(Descovy; Gilead Sciences, Foster City, CA) and the FTC/TDF study participants received emtricitabine 200 mg/tenofovir disoproxil fumarate 300 mg (Truvada; Gilead Sciences). The 33% regimen was defined as dosing on day 1 followed by skipped doses on days 2 and 3, repeated continuously; the 67% regimen as dosing on days 1 and 2 followed by a skipped dose on day 3, repeated continuously; and the 100% regimen as daily dosing throughout the 12-week period. The FTC/TDF study also included 33% and 67% regimens that skipped doses by weeks rather than days (the FTC/TAF study only included doses skipped by days, as described above). For example, half the FTC/TDF participants randomized to 33% dosing received daily dosing for one week followed by two weeks without dosing, repeated for 12 weeks and half the 67% group received two weeks of daily dosing followed by a week without dosing, repeated for 12 weeks. All doses were directly observed by study personnel either in person, by live video streaming, or via timestamped video recording. Blood was collected at baseline (pre-dose) and approximately four hours after the first dose (just the FTC/TAF study), then at weekly (FTC/TAF study) or biweekly (FTC/TDF study) intervals during dosing regimens and every three weeks during the washout period, without regard to time since the most recent dose (convenience sampling). For the FTC/TAF study, if a baseline sample was unavailable, the 4-h first-dose sample was used (n=4) and if a week 12 sample was unavailable the week 11 sample was used (n=13). For the FTC/TDF study, if a week 12 sample was not available the week 10 (n=3) or week 8 was used (n=1). Standard clinical laboratory assessments including complete blood count, comprehensive metabolic panel, phosphorus, and lipase were obtained at screening and every four weeks during active dosing.

### Dried Blood Spots

For each blood draw, 25 µL of whole blood was spotted five times onto a Whatman 903 protein saver card using a calibrated pipette.. All cards were air-dried at room temperature for a minimum of three hours (and up to overnight) and stored at −80°C until analysis.

#### DNA methylation

Genome-wide DNA methylation profiling was performed using the Infinium MethylationEPICV2 BeadChip (Illumina), which covers >850,000 CpG sites, following manufacturer protocols. Briefly, 500 ng of genomic DNA per sample was bisulfite-converted (Zymo EZ DNA Methylation kit) and hybridized to the EPIC array, with arrays scanned on an iScan instrument to produce raw intensity data. To pre-process the methylation data, we used the *minfi* pipeline^48^, and low quality samples were identified using the *qcfilter()* function from the ENmix package^49^, using default parameters. 100% of the original samples passed the QA/QC (p < 0.05) and were deemed to be high quality samples.

#### Epigenetic Clock Calculations

We examined three generations of epigenetic clocks: first-generation clocks that estimate chronological age (Horvath1, Horvath2, and Hannum clocks); second-generation clocks that predict mortality/morbidity risk (PhenoAge and GrimAge, including a principal-component version of GrimAge denoted PCGrimAge); and a third-generation clock known as DunedinPACE, which measures the pace of aging (years aged per chronological year). To improve robustness for longitudinal analysis, we used principal component-based versions of the first- and second-generation clocks (denoted “PC clocks”) wherever applicable. These PC clocks leverage principal components of DNAm data associated with the original clock algorithms, enhancing technical reliability and reducing noise in repeated measures. Published epigenetic clocks were calculated according to published methods from processed DNA methylation data. To calculate the principal component-based epigenetic clock for the Horvath multi-tissue clock, Hannum clock, DNAmPhenoAge clock, GrimAge clock, and telomere length we used the custom R script available via GitHub (https://github.com/MorganLevineLab/PC-Clocks). Non-principal component-based (non-PC) Horvath, Hannum, and DNAmPhenoAge epigenetic metrics were calculated using the *methyAge* function in the ENMix R package. The pace of aging clock, DunedinPACE, was calculated using the *PACEProjector* function from the DunedinPACE package available via GitHub (https://github.com/danbelsky/DunedinPACE). Supplemental Figure 1 shows cross-clock agreement with chronological age. We used a 12 cell immune deconvolution method to estimate cell type proportions^50^. Cell immune deconvolution results shown in Supplemental Figure 2.

#### Intracellular drug exposure and RepeatMasker-associated methylation analyses

Analyses were restricted to the subset of participants with paired DNA methylation data used for epigenetic age analyses and matched post-dose pharmacokinetic metadata. For each CpG, methylation change was defined as the post-treatment minus pre-treatment M-value (delta M). In the FTC/TAF study, to test whether interindividual variation in intracellular drug exposure was associated with methylation remodeling over the 12-week treatment period, we performed paired delta-epigenome-wide association analyses (delta-EWAS) separately for log-transformed PBMC tenofovir diphosphate (TFV-DP), emtricitabine triphosphate (FTC-TP), and tenofovir-diphosphate in dried blood spot (DBS) exposure measures. For each exposure, samples with non-missing post-dose values were retained, the exposure variable was standardized, and CpG-wise linear models were fit using limma. To identify repeat-associated loci, EPIC v2 hg38 CpG annotations were obtained from IlluminaHumanMethylationEPICv2anno.20a1.hg38, duplicate suffixes were removed to generate base CpG identifiers, and CpGs present in the EWAS results were converted to genomic ranges. These CpGs were intersected with hg38 RepeatMasker annotations obtained through AnnotationHub using findOverlaps after harmonizing chromosome naming conventions and restricting to standard chromosomes. Overlapping CpGs were then assigned to prespecified repeat groups, including SINE_Alu, SINE_MIR, LINE_L1, LINE_L2, LTR_ERV1, LTR_ERVK, LTR_ERVL, LTR_MaLR, DNA transposon, satellite, simple repeat, low-complexity, and an “Other” category. For each repeat class and exposure, we quantified the total number of CpGs, the proportion showing nominal association (*P* < 0.05), and the mean moderated *t*-statistic. Directional skew towards methylation gain or loss was assessed using a two-sided binomial test. Heatmaps display the number of nominally associated CpGs over the total number tested within each repeat class, with cell color indicating the mean *t*-statistic.

#### Statistical Analysis

All statistical analyses were performed in R (v4.5.1). For each epigenetic aging outcome including epigenetic clocks, system-specific “systems age” clocks, DunedinPACE and intrinsic capacity, DNA methylation–derived cell-type estimates, and epigenetic proxy biomarkers, we fit a linear mixed effects model: outcome ∼ timepoint + age + sex + (1|participant). Specifically, for each outcome *Y*, we fit a linear mixed-effects model where timepoint, age, and sex were modeled as fixed effects, and a participant-specific random intercept was included for participant identifier to model within-individual correlation. Models were fitted using lme4::lmer. For each outcome, we report the estimated *β*_1_with 95% confidence intervals (CI) and two-sided p-values. Results were visualized as forest plots showing the adjusted group difference (effect estimate) and 95% CI with a null reference at zero. Given the exploratory nature of the analyses, p-values were unadjusted.

#### Data Availability

The epigenetic data generated (DNAm beta values for EPIC arrays) and analyzed during this study will be made available in a public repository upon publication, with controlled access to protect participant privacy. Summary data for epigenetic clock measures and relevant clinical variables are provided in the Supplementary Information. The trial’s clinical data are available from the corresponding author on reasonable request, in accordance with institutional data sharing policies. Anonymized data will be shared by request from a qualified academic investigator for the sole purpose of replicating procedures and results presented in the article.

#### Code Availability

Epigenetic clock algorithms were applied using publicly available code or R packages as referenced above.

## Funding

This work was supported by the National Institutes of Health (U01 AI106499) and an investigator initiated study from Gilead Sciences (CO-US-412-3992). Its contents are solely the responsibility of the authors and do not necessarily represent the official views of the funders or NIH. *This publication was supported by the James B. Pendleton Charitable Trust and by the San Diego Center for AIDS Research (SD CFAR), an NIH-funded program (P30 AI036214), which is supported by the following NIH Institutes and Centers: NIAID, NCI, NHLBI, NIA, NICHD, NIDA, NIDCR, NIDDK, NIGMS, NIMH, NIMHD, FIC, and OAR*.

## Author contributions

Conceptualization: MJC and PA Methodology: MJC and PA Investigation: MJC and PA Supervision: MJC and PA Writing—original draft: MJC and PA Writing—review & editing: MJC and PA

## Competing interests

PLA receives research contracts from Gilead Sciences and Merck paid to his institution. AL receives research contracts from Gilead Sciences, Viiv Healthcare, and Merck paid to his institution. AL serves on a scientific review board for a research scholars program for Gilead Sciences. All other authors declare no other competing interests.

## Notes

### Clinical Trial

NCT02022657, NCT02962739

### Author Declarations

The FTC/TDF study was conducted at the San Francisco Department of Health and University of Colorado Anschutz Medical Campus, approved by the respective Institutional Review Boards, and was registered on ClinicalTrials.gov (NCT02022657); specimens were biobanked under a Colorado Multiple Institutional Review Board approved protocol. The FTC/TAF study was conducted at the University of Colorado Anschutz Medical Campus under a Colorado Multiple Institutional Review Board approved protocol and was registered on ClinicalTrials.gov (NCT02962739). All participants provided written informed consent prior to enrollment including consent for using stored samples for future genetic and other testing.

